# Outbreak of Kawasaki disease in children during COVID-19 pandemic: a prospective observational study in Paris, France

**DOI:** 10.1101/2020.05.10.20097394

**Authors:** Julie Toubiana, Clément Poirault, Alice Corsia, Fanny Bajolle, Jacques Fourgeaud, François Angoulvant, Agathe Debray, Romain Basmaci, Elodie Salvador, Sandra Biscardi, Pierre Frange, Martin Chalumeau, Jean-Laurent Casanova, Jérémie F. Cohen, Slimane Allali

## Abstract

**Background:** Acute clinical manifestations of SARS-CoV-2 infection are less frequent and less severe in children than in adults. However, recent observations raised concerns about potential post-viral severe inflammatory reactions in children infected with SARS-CoV-2.

**Methods:** We describe an outbreak of cases of Kawasaki disease (KD) admitted between April 27 and May 7, 2020, in the general paediatrics department of a university hospital in Paris, France. All children prospectively underwent nasopharyngeal swabs for SARS-CoV-2 RT-PCR, SARS-CoV-2 IgG serology testing, and echocardiography. The number of admissions for KD during the study period was compared to that observed since January 1, 2018, based on discharge codes, using Poisson regression.

**Results:** A total of 17 children were admitted for KD over an 11-day period, in contrast with a mean of 1.0 case per 2-week period over 2018-2019 (Poisson incidence rate ratio: 13.2 [95% confidence interval: 7.3-24.1], *p* <0.001). Their median age was 7.5 (range, 3.7-16.6) years, and 59% of patients originated from sub-Saharan Africa or Caribbean islands. Eleven patients presented with KD shock syndrome (KDSS) requiring intensive care support, and 12 had myocarditis. All children had marked gastrointestinal symptoms at the early stage of illness and high levels of inflammatory markers. Fourteen patients (82%) had evidence of recent SARS-CoV-2 infection (positive RT-PCR 7/17, positive IgG antibody detection 14/16). All patients received immunoglobulins and some received corticosteroids (5/17). The clinical outcome was favourable in all patients. Moderate coronary artery dilations were detected in 5 cases (29%) during hospitalisation.

**Conclusions:** The ongoing outbreak of KD in the Paris might be related to SARS-CoV2, and shows an unusually high proportion of children with gastrointestinal involvement, KDSS and African ancestry.

## INTRODUCTION

In children and adolescents, severe acute respiratory syndrome coronavirus 2 (SARS-CoV-2) infection is mostly responsible for mild respiratory symptoms, by contrast with severe forms reported in adults.^1 2^ An association between coronavirus disease 2019 (COVID-19) and late vasculitis manifestations has been increasingly suspected, especially in young asymptomatic patients, which may be due to post-viral immunological reactions.^3 4^

Kawasaki disease (KD) is the most common primary vasculitis in childhood, affecting predominantly medium and small-sized arteries.^5^ KD annual incidence is the highest in Japan, with more than 300 per 100 000 children < 4 years of age, compared to 25 per 100 000 children < 5 years of age in North America.^6 7^ One of the most severe complications of KD is coronary artery aneurysm.^7^ KD shock syndrome (KDSS) is a rare form of KD, frequently associated with myocarditis and requiring critical care support during the acute phase of illness.^8 9^ Although the aetiology of KD remains unclear, a role of a viral trigger on some genetically predisposed children has been hypothesised, as several viral respiratory agents have been associated with KD,^10^ including seasonal coronavirus in some,^11 12^ but not all, studies.^13 14^

Recently, nine cases of children with signs and symptoms consistent with KD and laboratory evidence of recent SARS-CoV-2 infection were reported in the United States and England.^15 16^ These reports included eight cases with hyperinflammatory syndrome and multiorgan involvement similar to what is seen in KDSS. Given the highly variable prevalence rates of SARS-CoV-2 infection in Europe, the possibility of an association between KD and positive testing for SARS-CoV-2 needs confirmation. Here we report a cluster of 17 patients with KD admitted to a Parisian university hospital’s paediatric department that occurred between April 27 and May 7, 2020, and evaluate potential temporal association with SARS-CoV-2 infection.

## PATIENTS AND METHODS

We included all children (age ≤ 18 years at the time of admission) diagnosed with KD and admitted in the department of general paediatrics and paediatric infectious diseases of Necker Hospital for Sick Children in Paris, France between April 27 and May 7, 2020. This university hospital serves as the regional reference centre for emerging infectious diseases in children. The study was approved by the Ethics Committee (Comité de Protection des Personnes Ouest IV, n° DC-2017-2987). All parents provided written informed consent.

The medical files of all patients were reviewed by using a standardised study-specific form to collect demographic and clinical data, laboratory test results, and imaging and echocardiographic findings. For the purposes of this study, the diagnosis of KD was based on the criteria of the American Heart Association for complete and incomplete KD,^7^ and the criteria proposed by Kanegaye *et al*. were used for the diagnosis of KDSS.^8^ All patients underwent nasopharyngeal swabs for SARS-CoV-2 RT-PCR (SARS-CoV2 R-GENE, Argene, bioMerieux), blood sampling for IgG SARS-CoV2 antibodies (SARS-CoV2 immunoassay, Abbott Core Laboratory) and laboratory tests, including inflammatory and cardiac markers. Standard cardiology workup included regular electrocardiograms and echocardiograms. A coronary artery dilation was defined as a Z-score of coronary artery diameter between 2 and <2.5 and aneurysm as a Z-score ≥ 2.5.^7^ Intravenous immunoglobulin (IVIG) resistance was defined as persistent or recrudescent fever at least 36 hours and <7 days after completion of first IVIG infusion.^7^

We compared the number of admissions for KD during the study period to that observed since January 1, 2018, based on discharge codes. We used Poisson regression to assess the change in the incidence of KD during the peak period, compared to all other two-week periods, after excluding the ongoing period (weeks 17-20, 2020). We described patients’ characteristics using medians and percentages. Differences between groups were assessed by the Mann-Whitney *U* test. Statistical analysis involved use of SPSS v21 (SPSS Inc., Chicago, IL).

## RESULTS

### Clinical features

A total of 17 children were admitted with a diagnosis of KD between April 27 and May 7, 2020. In comparison, the average number of children admitted in our department with a diagnosis of KD over 2018-2019 was 1.0 per 2-week period, for a Poisson incidence rate ratio of 13.2 (95% confidence interval: 7.3 to 24.1, *p* <0.001; **Figure 1**). Clinical features are presented in **Table 1**. The male/female ratio was 0.7. The median age at presentation was 7.5 years (range, 3.7-16.6), 59% had at least one parent originating from sub-Saharan Africa or Caribbean islands, and 12% from Asia. Patients had no relevant personal or familial history, and 14/17 (82%) were under the 97^th^ centile for body mass index. A diagnosis of complete KD was made for 8 (47%) children, while the others had incomplete KD. Among principal criteria for KD, polymorphous skin rash (76%) and bilateral bulbar conjunctival injection (76%) were the most frequent. All patients had gastrointestinal symptoms, occurring early in the course of illness before the onset of KD principal manifestations and consisting of acute abdominal pain, often associated with vomiting and diarrhoea (94%). Three had peritoneal effusion, with acute surgical abdomen in two cases. One of them underwent abdominal surgery for suspected appendicitis, revealing aseptic peritonitis. Irritability was common (65%), and 5 (29%) patients presented with headaches, confusion or meningeal irritation. Three underwent lumbar puncture, which revealed cerebrospinal fluid pleocytosis in one case. Among other acute manifestations of KD, 8 (47%) patients had pericardial effusions, and 3 (18%) had pleural effusion. Myocarditis was diagnosed in 12 (71%) patients, with left ventricular ejection fraction ranging between 10% and 57%. Two of them displayed significant electrocardiographic changes (increased QT interval and occasional ventricular arrhythmias or diffuse ST-segment elevation) not attributable to any QTc-prolonging drug.

**Table 1.** Clinical characteristics of included patients (N=17)

| Characteristics, (%) otherwise stated |  | Value |
| --- | --- | --- |
| Demographic features |  |  |
|  | Age, years; Median (range) | 7.5 (3.7-16.6) |
|  | Female | 10 (59) |
|  | Parents origin |  |
|  | sub-Saharan Africa/Caribbean islands | 20 (59) |
|  | Asia | 10 (29) |
|  | Europe | 4 (12) |
| KD principal clinical criteria |  |  |
| | Complete presentation (Fever > 4 days and ≥ 4 principal criteria) | 8 (47) |
|  | Lips and oral cavity changes | 12 (71) |
|  | Bilateral bulbar conjunctival injection | 13 (76) |
|  | Rash | 13 (76) |
|  | Extremities changes | 6 (35) |
|  | Cervical lymphadenopathy | 11 (65) |
|  | Days of fever before IVIG; Median (range) | 6 (1-13) |
| KD associated clinical features |  |  |
|  | Gastrointestinal symptoms | 17 (100) |
|  | Perineal or face desquamation | 4 (24) |
|  | Arthralgia | 2 (12) |
|  | Irritability | 11 (65) |
|  | Other neurological features | 5 (29) |
|  | Myocarditis | 12 (71) |
|  | Left ventricular ejection fraction rate; median (range)* | 38 (10-57) |
|  | Serous effusion | 8 (47) |

\*patients with myocarditis; IVIG: intravenous immunoglobulin. KD: Kawasaki disease.

**Figure 1.**
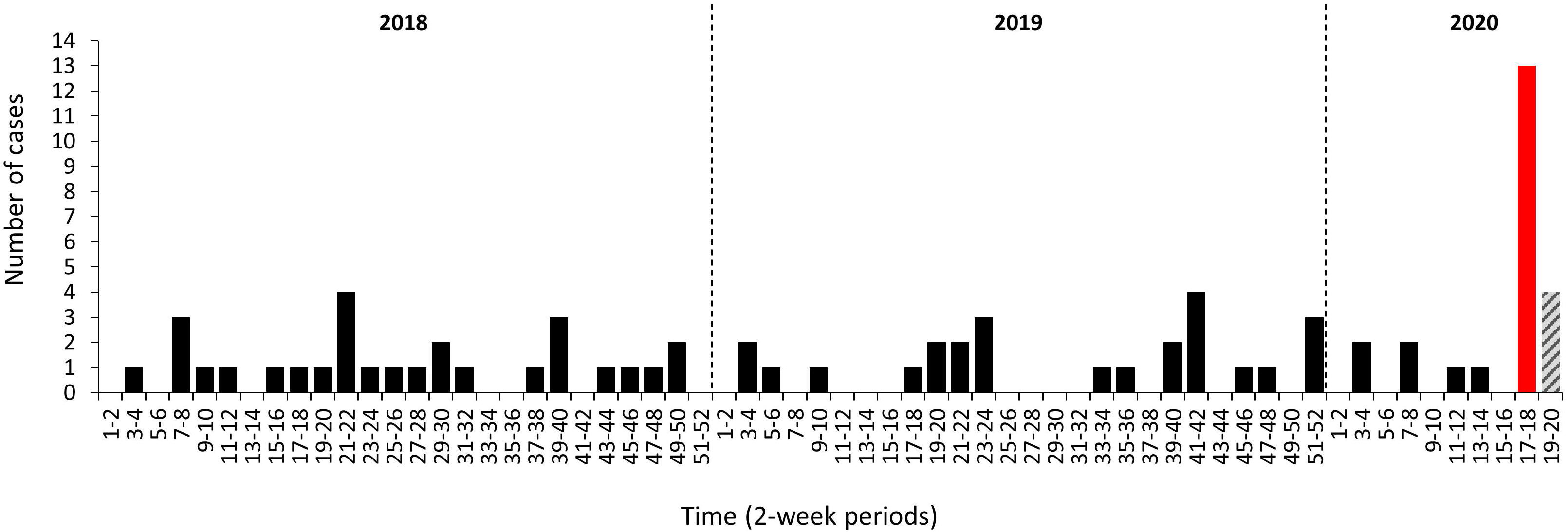
Number of admissions for Kawasaki disease in the department of general paediatrics of Necker Hospital for Sick children, by 2-week periods, 2018-2020 The grey zone corresponds to the ongoing 2-week period.

### Imaging and laboratory findings

Of the 13 patients who underwent chest imaging (X-ray or CT-scan), ground-glass opacities, local patchy shadowing, and interstitial abnormalities were present in 5 patients (38%) (**Table 2**). Echocardiography detected coronary arteries abnormalities in 8 patients (47%) after a median of 7.5 (range, 5-11) days of fever, which consisted of dilations for 5 patients (29%) and increased coronary visibility in three patients. No coronary aneurysm was identified. All patients had high inflammatory parameters, including leukocytosis with a predominance of neutrophils, and high levels of C-reactive protein (CRP), procalcitonin (PCT), and serum interleukin-6 (IL-6; **Table 2**). Anaemia was common with a median haemoglobin level reaching 8.1 (range, 5.3-12.2) g/dL. All patients had hyponatraemia (<135 mmol/L), and hypoalbuminaemia (<32 g/L) except one patient with mild KD. Transient kidney failure was observed in 8 (47%) of patients. Moderate elevation in serum alanine transaminases (ALT) and gammaglutamyl transpeptidase (GGT) occurred in 53% and 76% of patients, respectively, and GGT elevation occurred after several days of disease course. Lipase enzyme was elevated in 7 out of the 12 patients tested. D-dimers were elevated (>500 ng/mL) in 15/16 patients (94%). Elevated levels of troponin (>26 ng/mL) and B-type natriuretic peptide (BNP, >100 pg/mL) were found in 13/17 (76%) and 11/17 (65%) patients, respectively.

**Table 2.**
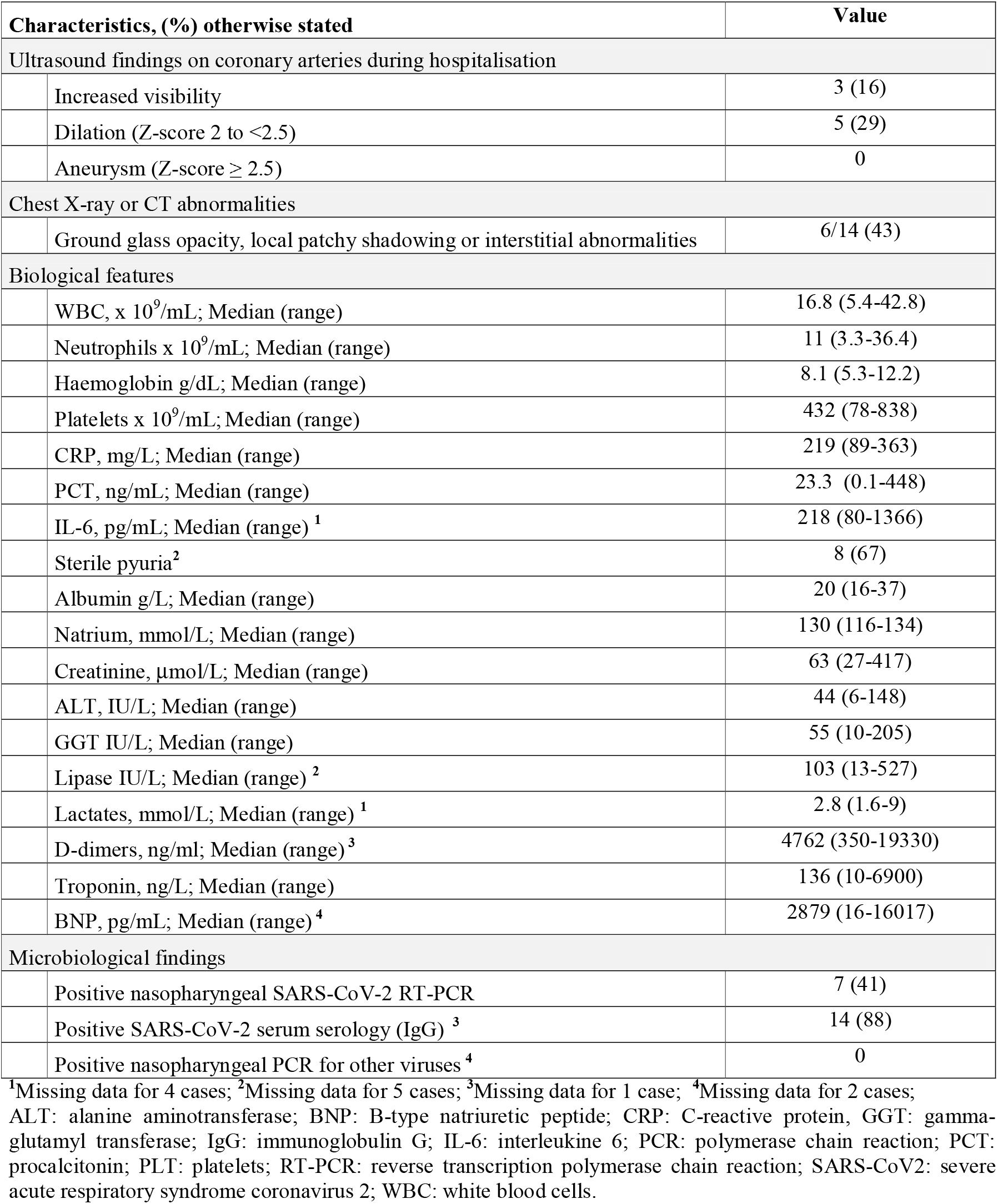
Imaging and laboratory findings of included patients (N=17)

| Characteristics, (%) otherwise stated |  | Value |
| --- | --- | --- |
| Ultrasound findings on coronary arteries during hospitalisation |  |  |
|  | Increased visibility | 3 (16) |
|  | Dilation (Z-score 2 to <2.5) | 5 (29) |
| | Aneurysm (Z-score $\geq 2.5$) | 0 |
| Chest X-ray or CT abnormalities |  |  |
|  | Ground glass opacity, local patchy shadowing or interstitial abnormalities | 6/14 (43) |
| Biological features |  |  |
|  | WBC, x 10<sup>9</sup>/mL; Median (range) | 16.8 (5.4-42.8) |
|  | Neutrophils x 10<sup>9</sup>/mL; Median (range) | 11 (3.3-36.4) |
|  | Haemoglobin g/dL; Median (range) | 8.1 (5.3-12.2) |
|  | Platelets x 10<sup>9</sup>/mL; Median (range) | 432 (78-838) |
|  | CRP, mg/L; Median (range) | 219 (89-363) |
|  | PCT, ng/mL; Median (range) | 23.3 (0.1-448) |
|  | IL-6, pg/mL; Median (range) <sup>1</sup> | 218 (80-1366) |
|  | Sterile pyuria<sup>2</sup> | 8 (67) |
|  | Albumin g/L; Median (range) | 20 (16-37) |
|  | Sodium, mmol/L; Median (range) | 130 (116-134) |
| | Creatinine, μmol/L; Median (range) | 63 (27-417) |
|  | ALT, IU/L; Median (range) | 44 (6-148) |
|  | GGT IU/L; Median (range) | 55 (10-205) |
|  | Lipase IU/L; Median (range) <sup>2</sup> | 103 (13-527) |
|  | Lactates, mmol/L; Median (range) <sup>1</sup> | 2.8 (1.6-9) |
|  | D-dimers, ng/ml; Median (range) <sup>3</sup> | 4762 (350-19330) |
|  | Troponin, ng/L; Median (range) | 136 (10-6900) |
|  | BNP, pg/mL; Median (range) <sup>4</sup> | 2879 (16-16017) |
| Microbiological findings |  |  |
|  | Positive nasopharyngeal SARS-CoV-2 RT-PCR | 7 (41) |
|  | Positive SARS-CoV-2 serum serology (IgG) <sup>3</sup> | 14 (88) |
|  | Positive nasopharyngeal PCR for other viruses <sup>4</sup> | 0 |

<sup>1</sup>Missing data for 4 cases; <sup>2</sup>Missing data for 5 cases; <sup>3</sup>Missing data for 1 case; <sup>4</sup>Missing data for 2 cases;
ALT: alanine aminotransferase; BNP: B-type natriuretic peptide; CRP: C-reactive protein, GGT: gamma-glutamyl transferase; IgG: immunoglobulin G; IL-6: interleukine 6; PCR: polymerase chain reaction; PCT: procalcitonin; PLT: platelets; RT-PCR: reverse transcription polymerase chain reaction; SARS-CoV2: severe acute respiratory syndrome coronavirus 2; WBC: white blood cells.

### Treatment and outcome

As first-line treatment, all 17 patients received high-dose IVIG (2 g/kg) after a median of 5 days of fever (range, 0-12) and low-dose aspirin (**Table 3**), and three patients received concomitant corticosteroids (2 to 10 mg/kg/day). Five (29%) had IVIG resistance and were treated with a second infusion of IVIG (2 g/kg), with the addition of corticosteroids (2 mg/kg/day) for three of these patients. Broad-spectrum antibiotics were administered to 14 patients (82%) until negative bacteriological results returned negative.

**Table 3.** Treatment and outcomes of included patients (N=17)

| **Characteristics, (%) otherwise stated** |  | **Value** |
| --- | --- | --- |
| **Treatment** |  |  |
|  | IVIG infusion | 17 (100) |
|  | IVIG retreatment | 5 (29) |
|  | Steroids | 5 (29) |
|  | Aspirin | 17 (100) |
|  | Broad-spectrum antibiotics | 14 (82) |
| **Outcome and serious complications** |  |  |
|  | Persistent fever at 36 hours after the end of the initial IVIG infusion | 5 (29) |
|  | Length of hospital stay, days; Median (range)* | 8 (5-17) |
|  | Hospitalisation in the intensive care unit | 13 (76) |
|  | Fluid resuscitation | 11 (65) |
|  | Vasoactive/inotropic agents | 10 (59) |
|  | Mechanical ventilation | 10 (59) |
|  | Death | 0 |

\* 5 patients not discharged; IVIG: intravenous immunoglobulin.

Thirteen patients (76%) were admitted to an intensive care unit (ICU) for management of haemodynamic instability. Eleven (65%) were considered as KDSS and received IV fluid resuscitation, together with vasoactive agents in 7 of them. Eight patients received inotropic agents in a context of myocarditis, and 9 needed respiratory support for management of cardiovascular compromise. ICU patients had higher systemic inflammation parameters, with a higher peak PCT level (52.4 ng/mL, range, 1.65-448) compared to non-ICU patients (1 ng/mL, range, 0.13-4.17; *p* = 0.002). Median ICU length of stay was 5.5 (range, 3-15) days. By May 7, 2020, 14 patients were discharged home after 8 (5-17) days of hospitalisation.

### Evidence of SARS-CoV-2 infection

In response to the SARS-Cov-2 pandemic, the French strategy of closing schools and the lockdown started on March 17, 2020. Since then, the 17 children reported here did not leave home for school, social gathering or travel, according to their parents. A recent history of viral-like symptoms was reported in 6 children: cough, coryza, fever < 48 hours, and for one patient, anosmia. The median duration between these preceding symptoms and onset of signs and symptoms of KD was 42 days (range, 18-79). History of recent contact with family members displaying viral-like symptoms was reported in 9 households: parents or grandparents (n=10), and siblings (n=3). Five of these family contacts had high suspicion of COVID-19 (ageusia, anosmia, suggestive findings on thoracic CT), of which one had a positive RT-PCR. The median interval between the reported contact and KD was 36 (range, 24-38) days. RT-PCR testing for SARS-CoV-2 in the 17 children presented here was positive for 7 (41%) (**Table 2**). All but one of them had no symptoms suggestive of Covid-19; one had anosmia that started 24 hours before KD symptoms. SARS-CoV-2 IgG antibodies were detected in 14 out of 16 (88%) tested patients. The two negative patients also had negative testing for RT-PCR SARS-CoV-2. These two patients presented with mild KD, with 5 and 6 days of fever before diagnosis, 3 and 4 principal KD criteria, and no myocarditis. Their PCT levels were < 1 ng/mL, and coronary dilatation was detected in one. Multiplex PCR targeting human respiratory syncytial viruses, seasonal coronaviruses, parainfluenza and influenza viruses, metapneumovirus and rhinovirus/enterovirus in nasopharyngeal swabs, was negative in all patients tested (n=16); serum PCR for adenovirus, EBV, CMV, HHV6, and parvovirus B19 were negative in the 9 tested children.

## DISCUSSION

### Interpretation

Compared to the two previous years, we found a 13-fold increased incidence in KD in children hospitalised in the general paediatrics department of a large university hospital centre in Paris. The temporal association with the onset of the SARS-CoV-2 epidemic in France and the results of RT-PCR and IgG testing in our patients suggest a causal link. Furthermore, all but one of our patients had no suggestive symptoms of acute Covid-19 disease and most had positive serum IgG responses, suggesting that the development of KD in these patients is more likely to be the result of a post-viral immunological reaction. Association between KD and viral respiratory infections has been previously suspected,^10 12 17^ especially rhinovirus/enterovirus and various viral agents including human coronaviruses.^10-12^ However, no difference in clinical presentation between infected and non-infected KD cases was previously reported.^10^

In our series, we recorded an over-representation of incomplete KD (53%), which might be explained by a high proportion of severe KD with myocarditis and KDSS, consistent with previous findings.^9^ Mild myocarditis is very common in the early phase of KD as demonstrated by cardiac biopsies and scintigraphy,^18 19^ and generally improves quickly as inflammation resolves.^18 20^ However, more severe myocarditis with decreased left ventricular contractility can sometimes occur, especially in the context of KDSS. KDSS is a rare complication affecting 1.5 to 7% of KD patients with a higher incidence in Western countries than in Asia.^9^ It appears to result from both myocardial dysfunction and decreased peripheral vascular resistance, usually requiring IV fluid resuscitation together with inotropic and vasoactive agent infusion in ICU.^8 21^ KDSS pathophysiology remains unclear. A high level of circulating pro-inflammatory cytokines may contribute to the distributive component of shock. Indeed, KDSS was previously found associated with high levels of IL-6, CRP and PCT.^9^ In our series, we observed very high levels of PCT, 8-fold higher than those recently reported in 27 KDSS patients.^9^ CRP and IL-6 levels were also high. This major pro-inflammatory state may reflect a particularly strong post-viral immunological reaction to SARS-CoV-2 as compared with other viral agents. Of note, a cytokine storm syndrome with elevated inflammatory markers such as IL-6 was described in adult COVID-19 patients,^22^ and has been associated with fatality.^23^

Besides inflammatory markers, clinical and biological features of our patients were often consistent with the diagnosis of KDSS. Indeed, older age, higher D-dimer, lower haemoglobin and albumin levels and more severe hyponatremia were previously found associated with KDSS.^9^ By contrast with the recent series of Riphagen et al,^16^ only 18% of our patients were above the 75^th^ percentile for weight, which does not support the hypothesis of overweight as a risk factor for KD after SARS-CoV-2 infection. IVIG resistance and coronary artery abnormalities were less frequent in our series than in previous studies.^8 9^ However, these results should be taken with caution as coronary artery abnormalities may appear later during follow-up. Gastrointestinal symptoms were also unusually common, affecting 100% of our patients. Miyake *et al*. reported intestinal pseudo-obstruction in only 2% of 310 patients.^24^ As previously described, other KD symptoms appeared in all cases after the intestinal ones, which may have led to diagnostic and therapeutic delays in some children.^25^ Suspected mechanisms involve intestinal ischemia, secondary to bowel vessel vasculitis. Rapid resolution of symptoms in all patients after IVIG supports this hypothesis. Pancreatitis, detected by elevated lipase level in 7 patients, may also reflect vasculitis.^26^ Hypoalbuminemia was severe and may partially be attributed to exudative enteropathy.

The observation of a higher rate of patients originating from sub-Saharan Africa and the Caribbean islands is consistent with recent findings reported by Riphagen *et al.,^16^* suggesting either adverse social and living conditions or genetic susceptibility. KD is rarely reported in sub-Saharan Africa, but it may be more common than previously thought.^27^ In the USA, a 2.5-fold higher incidence was reported in children of Asian than of European ancestry with an intermediary 1.5-fold risk for children of African ancestry.^28^ Besides, African Americans have been disproportionately hit by the COVID-19 pandemic, also suggesting an increased susceptibility to severe SARS-CoV-2 infection.^29 30^ Therefore, African countries where SARS-CoV-2 epidemic has spread may face a potentially large number of KD in children, and IVIG supply shortages should be anticipated in such settings. The absence of reported cases of KD associated with SARS-CoV2 infection in Asian countries where the SARS-CoV-2 epidemic started, and where the incidence of KD is the highest, is quite remarkable. Ethnic differences in the development of KDSS were previously reported, with a lower incidence rate in Asia than in Western countries.^8 9^ This warrants future studies investigating underlying genetic and immunologic mechanisms.

Our study has limitations; First, there is a potential recruitment bias that may have contributed to the significant rise in the number of KD patients hospitalised in our paediatric department. Our hospital is the referral centre for severe pediatric COVID-19 in the Paris area. Second, the low number of patients precluded in-depth comparisons of phenotypes with adequate statistical power.

### Conclusions

Our study documents an outbreak of KD in the Paris area and its association with recent SARS-CoV-2 infection. KD cases reported here have characteristics that differ from classic KD: this present form seems to be much more frequent among children of African ancestry, with predominant acute gastrointestinal manifestations, haemodynamic instability and myocarditis. These clinical findings should prompt a high degree of vigilance among primary care and emergency physicians, and preparedness in countries with a high proportion of children of African ancestry, while facing SARS-CoV-2 epidemic.

## Data Availability

The analyses as well as the anonymised database will be made available on reasonable request.

## ADDITIONAL STATEMENTS

### Contributors

Study conception: JT, SA, MC; Study design: JT, SA, MC; Data collection and curation: JT, CP; Investigation and resources: AC, FB, JF, FA, AD, RB, ES, SB, PF, JLC; Statistical analysis: JT, JFC; Writing the first draft: JT, SA, MC, JFC; Drafting the manuscript for important intellectual content: All authors.

### Dissemination to participants and related patient and public communities

Plans for dissemination include but are not limited to presentations in national and international scientific seminars.

## Acknowledgements

We are grateful to Prof. Christele Gras-Leguen (CHU Nantes, France) who was significantly involved in the ethical and regulatory considerations of this study. We also would like to thank Véronique Abadie, Christel Chalouhi, Mélissa Taylor, Morgane Le Gouez and Pierre Taupin, for their contribution to this study.

**ABBREVIATIONS**
CRP: C-reactive protein
CT: computed tomography
ICU: Inensive care unit
KD: Kawasaki Disease
KDSS: Kawasaki Disease Shock Syndrome
PCT: Procalcitonin

